# Restricted and repetitive behaviours are associated with noradrenergic alterations in neurodevelopmental disorders

**DOI:** 10.1101/2025.08.27.25334168

**Authors:** Sarah Al-Saoud, Emily S. Nichols, Abagail Hennessy, Vannessa Chen, Michelle Fang, Brian Krivoruk, Emma G. Duerden

**Author notes:** Correspondence to: Emma G. Duerden, PhD, Applied Psychology, Faculty of Education 1137 Western Rd, London, Ontario N6G 1G7.

## Abstract

Identifying early biomarkers of restricted and repetitive behaviours among children with neurodevelopmental disorders may support the development of targeted therapeutic interventions in both clinical and educational settings, ultimately improving their learning experiences and long-term developmental trajectories. The objectives of the study were to examine midbrain dopamine and norepinephrine integrity in children with and without neurodevelopmental disorders using neuromelanin-sensitive magnetic resonance imaging, and the association with restricted and repetitive behaviours. Participants were enrolled in the Child and Adolescent Brain Imaging Network (CABIN) or the Fetal Imaging and Neurodevelopment (FIND) studies. All participants completed MRI and behavioural assessments, including parent-reported RRBs measured with the Repetitive Behavior Scale–Revised. A total of 88 infants, children and adolescents (mean [SD] age, 9.47 [5.41] years; 46 males) were included. Age positively predicted neuromelanin-MRI signal intensity in the substantia nigra (β=.75, p<.001) and locus coeruleus (β=.38, p=0.007). Further, locus-coeruleus neuromelanin-MRI signal intensity was significantly higher in children with ASD and ADHD compared to TD peers (both, p<.007). Among children with autism, lower signal predicted more severe self-injurious behaviours (β=-.420, p=.006) and restricted interests (β=-.629, p<.001). Findings indicate a role for neuromelanin concentration in the locus coeruleus as a candidate biomarker for restricted and repetitive behaviours, particularly in children with autism. These behaviours may reflect reduced noradrenergic signalling and impaired capacity for arousal regulation and behavioural control in children with lower LC neuromelanin signal. Findings highlight the importance of targeting the noradrenergic system when developing therapeutic strategies. Pharmacological interventions aimed at modulating the noradrenergic system and school-based strategies supporting arousal regulation, behavioural flexibility, and cognitive control may help reduce symptom severity and improve functioning.

## Introduction

Neurodevelopmental disorders (NDDs) such as attention-deficit/hyperactivity disorder (ADHD) and autism spectrum disorder (ASD) comprise a diverse group of neuropsychiatric conditions with early onsets and produce impairments of adaptive, social, academic and/or occupational functioning^1^. Such impairments of functioning can in part be attributed to restricted and repetitive behaviours (RRBs), a broad and heterogeneous group of symptoms ranging from repetitive body movements to more complex behaviours, such as insistence on sameness or circumscribed interests. These maladaptive behaviours are mediated by alterations in dopaminergic (DA) and noradrenergic (NE) processes in the human midbrain^2^. Dysregulation of DA transmission can lead to an over-reliance on repetitive motor patterns, which may evolve into habitual actions^3^. In preclinical animal models, disruptions in DA signaling are associated with the emergence of repetitive behaviours^4^, and dysfunction of NE pathways are linked with behaviours that resemble the rigidity and compulsion often observed in ASD^5^. Recent advancements in neuromelanin magnetic resonance imaging (NM-MRI) provides a unique and non-invasive way of examining these pathophysiological mechanisms non-invasively without contrast agents in vivo among neurotypical and neurodiverse children by quantifying both DA and NE system levels in the substantia nigra (SN) and locus coeruleus (LC), respectively.

NM-MRI readily detects neuromelanin deposits that are preferentially found in catecholaminergic neurons of the midbrain, including DA neurons of the SN and NE neurons of the LC^6,7^. Neuromelanin is thought to reflect a chronological metric of cumulative metabolic waste because its concentration gradually accumulates in the SN and LC over the lifespan^6,8^. However, investigations probing neuromelanin accumulation over the lifespan have typically examined adults, omitting a key demographic group ranging from infancy to adolescence that may provide valuable insights into the early functioning of the DA and NE system. Previous research has provided support for using neuromelanin concentration in the SN and LC as proxy measures of DA and NE functioning in various neurodegenerative and neuropsychiatric disorders associated with aberrant movement, such as Parkinson’s disease^9,10^, schizophrenia^11,12^, and pediatric obsessive-compulsive disorder^13^, thereby warranting an examination of RRBs in children with NDDs using NM-MRI.

Growing evidence implicates dysfunction of the SN and LC in RRBs because DA circuits modulate motor processes^14^, and NE circuits modulate attention and inhibitory control^15,16^. Previous investigations have highlighted an association between motor control deficits and repetitive sensory-motor behaviours^17,18^, whereas restricted behaviours involving an undue fascination with objects, interests, or rituals may arise from a diminished ability to shift attentional focus^19,20^. Neuropharmacological research has illustrated the effectiveness of dopamine antagonists (e.g., risperidone) in decreasing sensory-motor behaviours such as rocking, pacing, hand-flapping^21^, and self-injurious behaviours such as head-banging and hair-pulling^22^, while adrenergic receptor agonists (e.g., methylphenidate, amphetamines) and antagonists (e.g., clonidine, guanfacine) have shown to improve response inhibition and attentional control among children with NDDs^23–26^. These findings suggest that DA and NE are key neuromodulators that mediate RRBs, however, further research is needed to elucidate their underlying neuropathophysiological mechanisms.

The present study examined midbrain DA and NE functioning in relation to RRBs among children with and without NDDs, as measured by NM-MRI signal intensity in the SN and LC, using in vivo imaging at high-field (3T) MRI. Our first aim was to quantify age-related changes in NM concentration in the SN and LC. We hypothesized that NM-MRI signal intensity in the SN and LC would increase with chronological age, given corresponding findings from previous investigations involving adults^6^. Our second aim was to characterize RRBs in the pediatric sample using NM-MRI as a proxy measure of DA and NE functioning. We hypothesized that alterations in LC- and SN-NM-MRI signal intensity would predict more severe RRBs, given evidence from pre-clinical research that implicates the DA and NE system in RRBs.

## Method

This study was approved by the Health Sciences Research Ethics Board (HSREB) at the Western University. All participants provided written informed consent or verbal assent. Written parental consent was obtained on behalf of all participants under 18 years old.

### Participants

A community-based sample of 75 typically developing (TD) and neurodiverse (ADHD and ASD) infants, children, and adolescents were recruited through the Child and Adolescent Brain Imaging Network (CABIN), an ongoing developmental neuroimaging initiative at Western University in London, Ontario. The CABIN cohort included a small number of infants (aged 6 months and older), enrolled due to familial risk (e.g., having an older sibling with a diagnosis) or early emerging developmental concerns reported by caregivers. Additionally, a sample of 11 typically developing infants was recruited from the community through the Fetal Imaging and Neurodevelopment (FIND) study, which includes cross-sectional and longitudinal data collected between August 2021 and September 2023. Infants in the FIND cohort were included as typically-developing controls. Participants from both cohorts were recruited using flyers displayed in the community and through advertisements on social media. In the CABIN cohort, the participants in the NDD group over the age of 4 had a primary diagnosis of ADHD or ASD, that was provided by a developmental paediatrician, psychiatrist or clinical psychologist. For TD participants in the CABIN and FIND cohorts, parents indicated via self-report that their infants/children had not been diagnosed with any psychological disorder(s). Inclusion criteria for infants recruited from FIND were singleton pregnancy, maternal age ≥ 18 years and age-appropriate fetal growth, whereas exclusion criteria included antenatal exposure to medications or illicit drug use, and contraindication to safety while undergoing MRI. The participant demographics are listed in Table 1.

**Table 1.** Participant demographics.

|  | TD | ADHD | ASD | Total |
| --- | --- | --- | --- | --- |
| CABIN Cohort |  |  |  |  |
| N | 28 | 36 | 11 | 75 |
| Age (years), mean, (SD) | 9.29(3.99) | 12.33(4.18) | 10.73(4.71) | 10.96(4.36) |
| Sex (males), n(%) | 13(46) | 19(53) | 6(55) | 50.7(38) |
| FSIQ (n) | 106(18) | 93(13) | 89(16) | 98(16) |
| FIND Cohort |  |  |  |  |
| N | 13<sup>a</sup> | - | - | 13 |
| Age (years), mean(SD) | 0.83(0.28) | - | - | 0.83(0.28) |
| Sex (males), n(%) | 8(62) | - | - | 8(62) |
| ASQ |  |  |  |  |
| Communication | 56.53(19.1) | - | - |  |
| Personal-social | 50.38(16.6) |  |  |  |
| Fine Motor | 35.77(20.6) |  |  |  |
| Gross Motor | 74.23(16.2) |  |  |  |
| Total |  |  |  |  |
| N | 41 | 32 | 11 | 88 |
| Age (years), mean, (SD) | 7.24(4.96) | 12.63(4.26) | 10.73(4.71) | 9.69(5.27) |
| Sex (males), n(%) | 23(51) | 18(56.3) | 6(54.5) | 48(55) |
| Abbreviations: TD, typically developing; ADHD, attention-deficit/hyperactivity disorder; ASD, autism spectrum disorder; FSIQ, Full Scale Intellectual Quotient; NA, not available. |  |  |  |  |
| <sup>a</sup> Infants (≤12 months old) were coded as typically developing (TD) because they do not meet the minimum age requirement for a neurodevelopmental disorder diagnosis. |  |  |  |  |

Altogether with both CABIN and FIND cohorts, a total sample of 88 participants between the ages of 3 months and 21 years (46 boys and 42 girls, M_age_ = 9.47 years, SD = 5.41 years) were scanned on the same NM-MRI protocol at 3T. Infants were assessed for developmental delays using the Ages and Stages Questionnaire (ASQ) and mean scores were in typically-developing ranges (Table 1). A representative sample of 26 participants in the CABIN cohort also underwent psychoeducational testing and were determined to have average cognitive functioning (i.e., Full Scale Intellectual Quotient ≥ 85), as measured using the Weschler Intelligence Scale for Children, Fifth Edition (WISC-V)^27^.

### Procedure

Imaging data including anatomical and NM-MRI scans were collected at Robarts Research Institute at Western University. Participants in the CABIN cohort were permitted to watch TV shows or movies during the MRI scan. Participants in the FIND cohort were scanned during natural sleep. Parent-report questionnaires were completed on-site or at home.

### Measures

RRBs were measured in participants 6 years and older using the Repetitive Behaviors Scale - Revised (RBS-R), which consists of 43-items across five subscales, including: Stereotypic Behavior (i.e., repeated movements with no obvious purpose), Ritualistic/Sameness Behaviour (i.e., resistant to change; fixed patterns of behaviour), Self-Injurious Behaviour (i.e., repeated movements/actions directed toward the body that have the potential to cause injury), Compulsive Behaviour (i.e., behavior that is repeated and performed according to a rule or involves things being done in a particular manner), and Restricted Interests (i.e., inflexible range of focus; intense or unusual interests or activities)^28,29^. The five-factor model proposed by Lam and Aman^29^, mapping onto the five subscales of the RBS-R, has been validated in children with ASD^30,31^. Interrater reliability data indicates that the RBS-R performs well in outpatient settinga, and measures of internal consistency are high for the five-factor solution^29^. The factor structure of the RBS-R is considered invariant across diagnoses of ASD and ADHD, which allows for empirically-validated group comparisons^32^.

The Repetitive Behavior Scale for Early Childhood (RBS-EC), which incorporates developmentally appropriate items for young children, was used to measure RRBs in participants 4-6-years-old^33^. The RBS-EC consists of 34 items across four subscales, including: Repetitive Motor, Ritual and Routine, Restricted Interests and Behavior, and Self-Directed Behavior. Notably, items reflecting adult-oriented compulsive behaviours from the RBS-R were excluded^34^. An examination of the RBS-EC in a community sample provides evidence of strong psychometric properties and concurrent criterion validity with the RBS-R^33^.

### Neuromelanin-Sensitive Magnetic Resonance Imaging MRI Acquisition

Magnetic resonance images were acquired from all participants on a 3T Prisma Fit scanner (Siemens, Erlangen, Germany). CABIN participants were scanned with a 32-channel head coil at Western University’s Centre for Functional and Metabolic Mapping. The FIND participants were scanned with a custom-built infant 32-channel head coil^35^. No differences in NM-signal (LC,SN) were evident between the coil types in the infants scanned for CABIN and FIND (LC p=.27 and SN p=.63). To measure NM-MRI signal concentration in all participants, a modified three-dimensional (3D) gradient-recalled echo with magnetization-transfer pulse (GRE-MT) NM-MRI image was acquired with the following parameters: repetition time (TR)=54ms; echo time (TE)=7.34ms; flip angle=16°; FoV=192×192; matrix=320×320; slice thickness=3mm; acquisition time=5:12min. A T1-weighted (T1w) image was acquired for NM-MRI image processing using a 3D magnetization prepared rapid acquisition gradient echo (MPRAGE) sequence with the following parameters: spatial resolution=1×1×1mm^3^; TR=2,300ms; TE=2.88ms; inversion time (TI)=900ms; flip angle=9°; acquisition time=5:00min; FoV=240×256; matrix=240×256; slice thickness=1mm; GRAPPA=2. Immediately upon acquisition, the NM-MRI images were visually inspected for artifacts affecting the midbrain due to participant motion.

### NM-MRI Preprocessing

The NM-MRI scans were preprocessed using Statistical Parametric Mapping (SPM12)^36^ in MATLAB (vR2022b) to allow for voxelwise analyses in standardized MNI (Montreal Neurological Institute) space. The same preprocessing pipeline was applied to all participants including data from infants, children, and adolescents. Given developmental differences in brain morphology and myelination, infant scans were visually inspected at each preprocessing stage to ensure anatomical alignment and quality. The steps of the preprocessing pipeline were as follows: 1) NM-MRI images were realigned to correct for motion using ‘SPM-Realign: Estimate and Reslice’; 2) The realigned and estimated NM-MRI images were averaged using ‘SPM-ImCalc’; 3) The averaged NM-MRI image was then co-registered to the T1w image using ‘SPM-Coregister: Estimate and Reslice’; 4) The realigned T1w image was segmented into different tissue types and spatially normalized to a standard MNI template using DARTEL^37^; 5) The spatially normalized NM-MRI image was spatially smoothed with a 1mm full-width-at-half-maximum 3D Gaussian kernel using ‘SPM-Smooth’; 6) The smoothed NM-MRI image was visually inspected relative to the template using ‘SPM-Check Reg.’

### Manual Segmentation

ITK-Snap (v. 3.8.0, www.itksnap.org)^38^ was used to manually segment the SN and LC in native MRI space. Additionally, the crus cerebri (CC) and pontine tegmentum (PT) were manually segmented as reference regions for subsequent contrast-to-noise ratio (CNR) analyses, as white-matter tracts are known to have minimal neuromelanin content^39^. The manual segmentation protocol followed previously described methods^39–41^ with reference to an anatomical MRI atlas of the mesencephalon^42^. Sample masks of the SN, LC, CC, and PT in a representative participant can be found in Figure 1.

**Figure 1.**
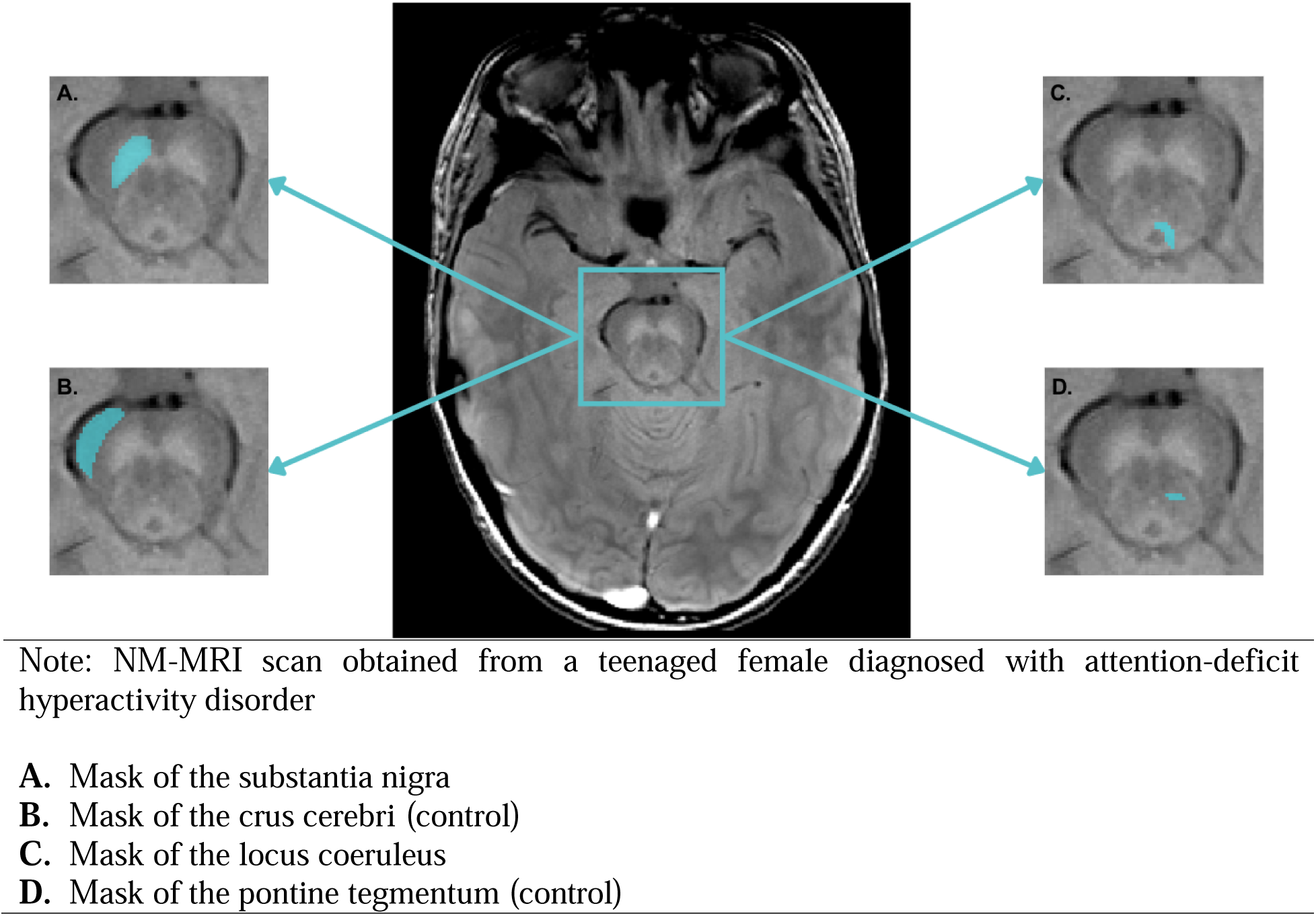
Manual segmentations in a representative participant.

### Contrast-to-Noise Ratio Calculations

Manual tracking-based segmentations methods were used to quantify the contrast between the signal of interest and background noise, which is particularly important when examining the SN and LC because they are small structures located in areas of the brain with complex anatomy^43^. A CNR value corresponding to a single ROI is the most widely used metric in NM-MRI studies, and its formula has been adapted from prior work: [(signal – reference)/reference*100)], yielding a metric of neuromelanin percent signal change relative to reference regions^39,13^. SN and LC masks were averaged bilaterally to account for intraindividual differences.

### Statistical Analysis

Statistical analyses were performed using the IBM Statistical Package for Social Sciences (SPSS, Version 29). One-way analyses of variance (ANOVAs) were conducted to examine demographic differences between diagnostic groups (TD, ASD, ADHD). Generalized linear models were run to examine NM-MRI signal intensity related to age and RRBs (stereotyped behaviours, self injury, compulsive behaviours, ritualistic behaviours, restricted interests).

To address our first aim, age was the independent variable and NM-MRI signal intensity in the SN and LC were the dependent variables (two separate models), adjusting for sex with diagnostic group as a factor. Power analysis indicated 77 participants were needed to detect a medium effect (f² = 0.15) at 80% power. Bonferroni corrections were used for multiple comparisons (p=0.05/2).

To address our second aim, RRBs were the dependent variables (five separate models) and the interaction between group and NM-MRI signal intensity in the SN and LC were the independent variables, adjusting for sex and age with diagnostic group as a factor. The alpha level was set at p=0.01(p=0.05/5).

## Results

### Participant Demographics

88 infants, children, and adolescents, including 47 participants with NDDs (ADHD n=36 and ASD n=11) and 41 TD participants took part in the present study (Table 1). Despite commonly reported differences in diagnosis rates across sex^44,45^, participant sex did not significantly differ between groups (X^2^=0.05, p=.978).

### Restricted and Repetitive Behaviours and Diagnostic Group

Nine participants did not complete the questionnaires including children with ADHD (n=3), ASD (n=2), and TD children (n=4). Stereotypic behaviour significantly differed between diagnostic groups (F(2,63)=3.89, p=.026), such that children with ASD reported greater stereotypic behaviour than TD children (p=.021, 95% C.I. = [0.82, 13.14]). Further, greater ritualistic behaviour was observed among children with NDDs (F(2,63)=9.23, p<.001), such that children with ASD reported higher ritualistic behaviour than children with ADHD (p=.006, 95% C.I. = [2.60, 19.45]) and TD children (p<.001, 95% C.I. = [6.28, 22.11]). Restricted interests also significantly differed between groups (F(2,63)=7.62,p=.001), such that children with ASD reported higher restricted interests than children with ADHD (p=.003, 95% C.I.=[1.79, 10.61]) and TD children (p<.001, 95% C.I.=[2.42, 11.28]). Total RRBs were found to be significantly higher among children with NDDs (F(2,63)=6.93, p=.002), and children with ASD demonstrated greater RRBs than children with ADHD (p=.033, 95% C.I.=[1.42, 45.90]) and TD children (p=.001, 95% C.I. = [11.35, 56.01], Table 2).

**Table 2.** RRB Scores by Diagnosis.

|  | TD | ADHD | ASD | Missing | Total |
| --- | --- | --- | --- | --- | --- |
| CABIN Cohort |  |  |  |  |  |
| N | 28 | 29 | 9 | 9 | 66 |
| Age (years), mean, (SD) | 9.04(3.78) | 12.69(4.40) | 10.56(5.25) | 11.78(2.81) | 10.85(4.54) |
| Sex (males), n(%) | 14(50.0) | 15(51.72) | 55.55(5) | 4(44.4) | 34(51.51) |
| RRB Scores, mean (SD) |  |  |  |  |  |
| Stereotypic Behaviour | 3.08(1.11) | 6.00(1.26) | 9.71(4.20) | NA | 5.26(6.82) |
| Self-Injurious Behaviour | 1.17(0.56) | 3.63(0.86) | 2.71(1.38) | NA | 2.77(4.05) |
| Compulsive Behaviour | 1.50(0.57) | 3.22(0.93) | 5.00(1.75) | NA | 2.86(4.04) |
| Ritualistic Behaviour | 4.04(1.31) | 8.44(2.13) | 18.29(3.97) | NA | 8.45(9.99) |
| Restricted Interests | 1.96(0.74) | 2.70(0.64) | 7.29(3.84) | NA | 3.26(5.16) |
| Total RRBs | 11.54(3.76) | 24.00(5.23) | 42.14(12.34) | NA | 22.42(25.76) |
| Abbreviations: TD, typically developing; ADHD, attention-deficit/hyperactivity disorder; ASD, autism spectrum disorder; RRB, restricted and repetitive behaviours; NA, not available. |  |  |  |  |  |

**Table 3.** NM-MRI Signal Intensity in the SN and LC by Diagnosis.

|  | TD | ADHD | ASD | Total |
| --- | --- | --- | --- | --- |
| CABIN Cohort |  |  |  |  |
| N | 28 | 36 | 11 | 75 |
| NM-MRI %, mean (SD) |  |  |  |  |
| <sup>a</sup> Substantia Nigra | 13.33(4.46) | 15.35(3.11) | 14.61(3.02) | 14.49(3.74) |
| <sup>b</sup> Locus Coeruleus | 10.47(2.77) | 15.79(7.45) | 17.29(8.35) | 14.03(6.81) |
| FIND Cohort |  |  |  |  |
| N | 13 | NA | NA | 13 |
| NM-MRI %, mean (SD) |  |  |  |  |
| <sup>a</sup> Substantia Nigra | 2.49(2.08) | NA | NA | 2.49(2.08) |
| <sup>b</sup> Locus Coeruleus | 5.32(2.98) | NA | NA | 5.32(2.98) |
| Total |  |  |  |  |
| N | 41 | 32 | 11 | 86 |
| NM-MRI %, mean (SD) |  |  |  |  |
| <sup>a</sup> Substantia Nigra | 10.60(6.11) | 15.53(3.17) | 14.61(3.02) | 12.95(5.37) |
| <sup>b</sup> Locus Coeruleus | 9.07(3.57) | 16.50(7.59) | 17.30(8.36) | 12.89(7.09) |
| Abbreviations: TD, typically developing; ADHD, attention-deficit/hyperactivity disorder; ASD, autism spectrum disorder; SN, substantia nigra; LC, locus coeruleus; NM-MRI, neuromelanin-sensitive magnetic resonance imaging; NA, not available. |  |  |  |  |
| <sup>a</sup> Percent signal change calculated relative to the crus cerebri (control) |  |  |  |  |
| <sup>b</sup> Percent signal change calculated relative to the pontine tegmentum (control) |  |  |  |  |

### NM-MRI Percent Signal Change, Age, and RRBs across Diagnostic Groups

Participant age significantly predicted SN NM-MRI signal intensity (β=.75, p<.001, Fig. 2A); however, diagnostic group differences were not evident (p=.27, Fig. 2B). Findings were maintained in the CABIN cohort alone with age being predictive of SN NM-MRI signal (β=.48, p<.001), with no group differences (p=.73). As no group differences in SN NM-MRI signal were evident, no further analyses were performed.

**Figure 2.**
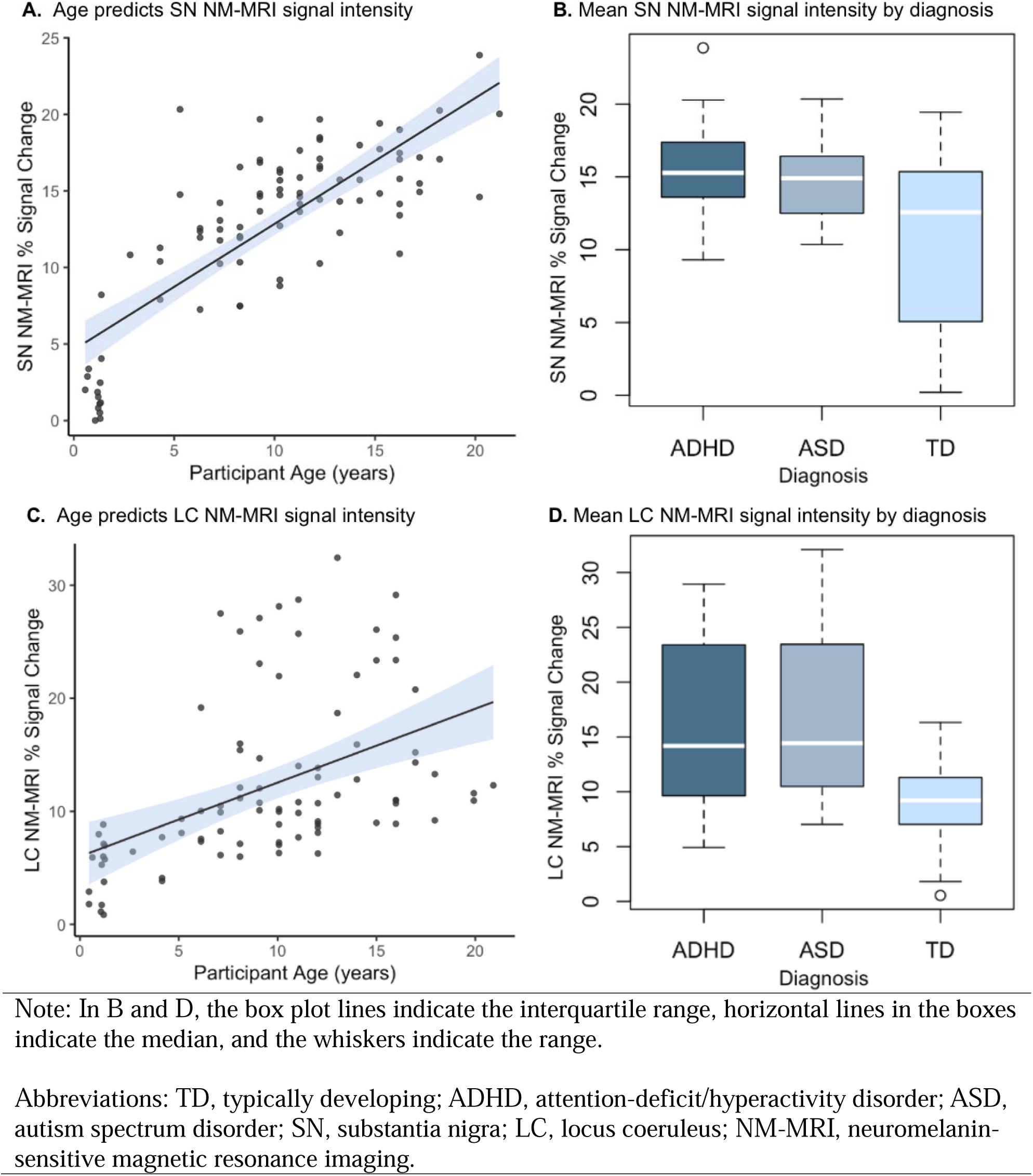
Neuromelanin concentrations in the substantia nigra and locus coeruleus.

Age also predicted LC-NM MRI signal intensity (β=.38, p=0.007; Fig. 2C), with significant group differences (p<.001; Fig. 2C) seen between participants with ASD (p=.003) and ADHD (p=.007) relative to their TD peers (Fig. 2D, Bonferroni corrected). In the CABIN cohort, group differences in LC NM-MRI signal were evident (p=.003), but age effects were no longer present (β=0.27, p=.12).

LC NM-MRI signal intensity significantly predicted more severe self-injurious behaviours (β=-.420, p=.006) and restricted interests (β=-.629, p<.001) in the ASD group only, based on an interaction analysis, adjusting for age and sex. Group effects were evident in both models (p<0.01).

## Discussion

This study demonstrates an association between RRBs and NE dysregulation in children with and without NDDs using NM MRI. Alterations in NE system functioning was associated with more severe RRBs in children with ASD and ADHD. Findings support a growing body of evidence implicating the LC-NE system in behavioural rigidity and arousal dysregulation across NDDs. Findings further provide evidence to suggest that neuromelanin deposits in the SN and LC increase with chronological age in infancy and early childhood. Developmental patterns align with prior histological and imaging research indicating progressive catecholaminergic system maturation during early life. Inclusion of infants and young children in both NDD and TD groups offers preliminary insight into individual differences in brainstem neurobiology across early developmental stages.

### Age Predicts NM-MRI Signal Intensity

Age positively predicted NM-MRI signal intensity in the SN and LC and corresponds with a larger body of evidence indicating that NM signal can be detected in early childhood and dramatically increases into adolescence^6,8^. In post-mortem research, significant accumulations of NM in the SN from infancy through old age was previously reported in a sample aged 1-97 years old^46^. This trajectory corresponds with a developmentally normative shift towards increased DA and NE system functioning^47,48^.

### Restricted and Repetitive Behaviours Differ by Diagnosis

Consistent with previous literature, RRBs were most severe among children with ASD including stereotypic, ritualistic, and restricted interest behaviours compared to children with ADHD and TD children^1^. Several factors contribute to RRBs in children with ASD, including heightened sensory sensitivities, reduced behavioural flexibility, and altered emotional regulation^49^. RRBs may serve as self-regulatory strategies in ASD. Social disengagement may also play a role, as RRBs can interfere with opportunities for reciprocal interaction and learning^50^. Evidence suggests that RRBs in ADHD may stem from broader difficulties in motor control, impulsivity and emotional regulation^51^.

### RRBs and LC-NE System processing

Our findings of elevated LC neuromelanin signal in children with ASD and ADHD aligns with pediatric OCD research showing increased midbrain neuromelanin signal, suggesting dopaminergic hyperactivity despite negative associations with symptom severity^13^. Although children with ASD demonstrated higher LC neuromelanin signal on average, within the ASD group, children with lower LC signal exhibited more severe restricted and self-injurious behaviours. Findings suggest that a relative reduction in LC integrity or noradrenergic function within an already dysregulated system may impair behavioural flexibility and increase vulnerability to repetitive or injurious behaviours, potentially due to the role of the NE system in response inhibition and attentional control^16,23^.

Contrary to our hypothesis, SN neuromelanin signal did not differ between groups. However, strong age-related effects were observed in SN signal, consistent with known developmental increases in neuromelanin concentration during childhood and adolescence^6,8^. Age-related variability in the signal may have reduced the ability to detect group-level differences, underscoring the importance of accounting for developmental stage in future work.

### Limitations

MR scanning of young children limited the inclusion of non-verbal participants and participants with pronounced motor activities who can struggle to remain still for extended periods, which limits the generalizability of our findings. Additionally, NM-MRI provides a proxy measure of DA and NE system functioning, meaning it cannot directly measure neurotransmitter synthesis and transmission. Despite this limitation, NM-MRI is appropriate for conducting developmental research as it does not require exposure to contrast agents.

## Conclusions

This study provides evidence for alterations in NM concentration in the LC, as measured using NM-MRI, as a candidate biomarker for RRBs, highlighting the importance of the targeting the NE system when developing therapeutic interventions. While NM-MRI offers a non-invasive method for examining brainstem catecholaminergic systems *in vivo*, further research is needed to determine its utility in identifying clinically meaningful subgroups or predicting treatment responses. NM-MRI may aid in informing school-based or behavioural interventional therapies. Findings further highlight the LC-NE system as a promising target for treating self-injurious behaviours using pharmacological or neurosurgical methods.

## Data Availability

All data produced in the present study are available upon reasonable request to the authors.

